# Multicohort assessment of plasma metabolomic measurements across the atherosclerosis continuum

**DOI:** 10.64898/2026.01.15.26344196

**Authors:** Sergi Sayols-Baixeras, Kamalita Pertiwi, Koen F. Dekkers, Tania Sharma, Luka Marko Rašo, Mario Delgado-Velandia, Gabriel Baldanzi, Ulf Hammar, Germán D. Carrasquilla, Stefan Gustafsson, Kim Kultima, Henrik Carlsson, Ida Erngren, Tammy Y. N. Tong, Alexis Elbaz, Adam S. Butterworth, Sölve Elmståhl, Kristian Hveem, Peter M. Nilsson, Markus Perola, Hemmo Sipsma, Birgit Simell, Bjørn O. Åsvold, Thomas Engstrøm, Akiko Maehara, Michael Maeng, Gregg W. Stone, Göran Bergström, Jan Borén, Clemens Wittenbecher, Johan Ärnlöv, Lars Lind, Gunnar Engström, Johan Sundström, J. Gustav Smith, David Erlinge, Tove Fall

## Abstract

**Aims:** Atherosclerosis develops over many years, and its underlying mechanisms are still not fully understood. Studying plasma metabolites at different stages of the disease may help identify biomarkers that clarify how it develops and improve early risk assessment.

**Methods:** We performed untargeted plasma metabolomics with ultra-performance liquid chromatography–mass spectrometry in 8,146 participants without cardiovascular disease from the population-based SCAPIS cohort. We examined how 1,171 circulating metabolites were associated with subclinical coronary atherosclerosis. Atherosclerosis burden was measured using coronary computed tomography angiography and quantified with the segment involvement score, and associations were analyzed using multivariable models. Associated metabolites were then evaluated in independent cohorts with phenotypes representing later stages of the atherosclerotic disease continuum: myocardial infarction within six months (MIMI, n=2,018), and coronary plaque burden and vulnerability in myocardial infarction survivors (PROSPECT II, n=898).

**Results:** Twelve metabolites, including phosphate, malate, sphingomyelins, and amino acids were robustly associated with subclinical coronary atherosclerosis after adjustment for traditional risk factors and additional clinical variables. Notably, malate, N-acetyl-isoputreanine and a nucleoside derivative were positively associated with subclinical coronary atherosclerosis and with myocardial infarctions. Sphingomyelins showed inverse associations with subclinical atherosclerosis, myocardial infarction, and the presence of vulnerable plaques.

**Conclusions:** These findings reveal a metabolomic signature across the atherosclerosis continuum, highlighting candidate biomarkers that may enhance understanding of disease mechanisms and aid risk stratification.

**Translational Perspective:** Coronary atherosclerosis often remains silent until advanced stages, highlighting the need for early detection of individuals at high-risk. Using plasma metabolomics, we identified twelve metabolites associated with subclinical coronary atherosclerosis in participants without prior cardiovascular disease. Several metabolties, including malate, phosphate, specific sphingomyelins, and a nucleoside-like compound, were consistently associated across the atherosclerosis continuum in independent cohorts. These findings underscored potential biomarkers that may advance our understanding of cardiovascular disease and improve cardiovascular risk assessment. However, further studies are needed to determine their clinical utility.

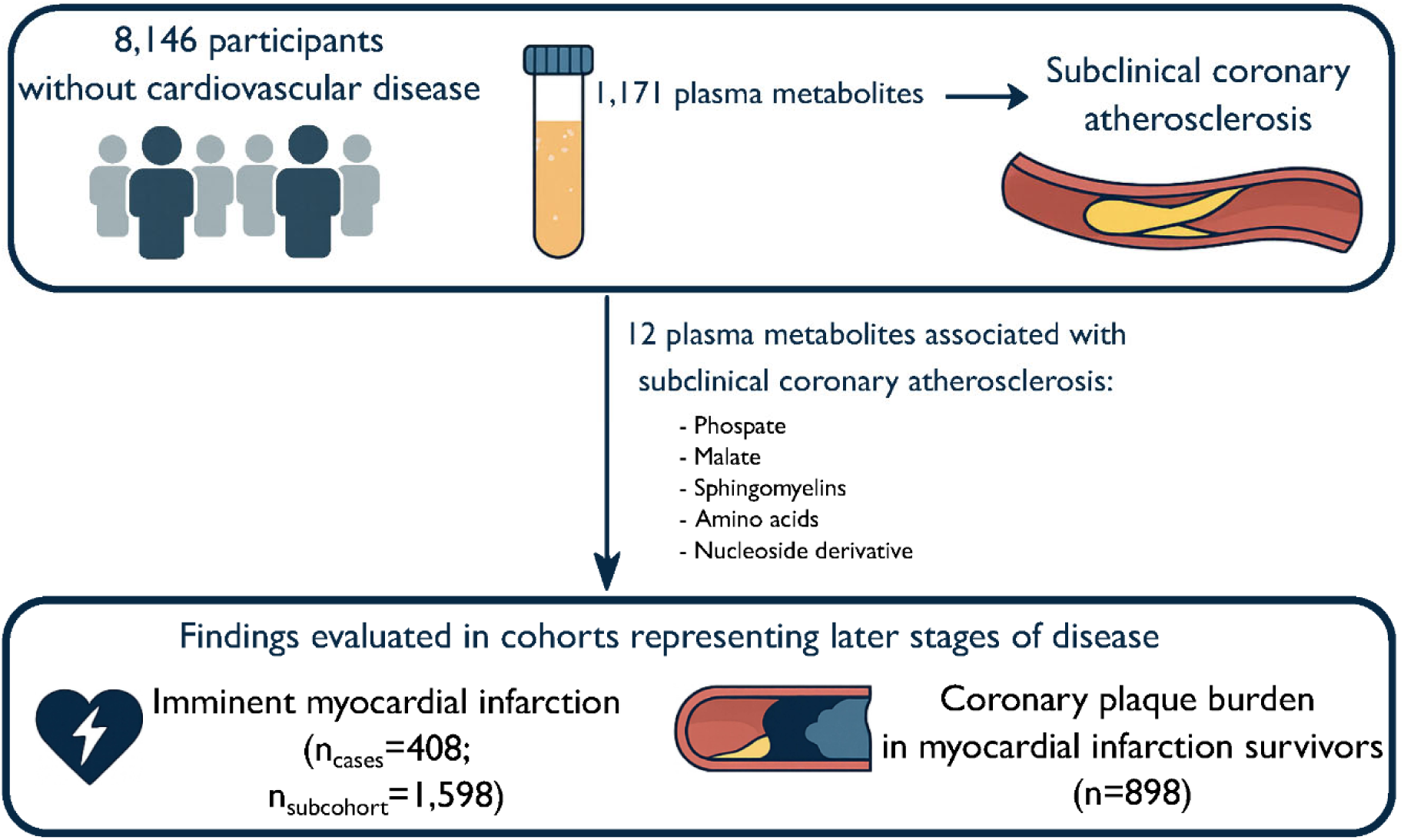
Graphic element.

## Introduction

Atherosclerosis is the leading cause of myocardial infarction, stroke, and peripheral artery disease worldwide.^1^ Its multifactorial pathogenesis is not fully understood but the disease usually progresses for decades, both before and after the onset of symptomatic cardiovascular events. Atherosclerotic lesions can be detected through advanced imaging techniques like coronary computed tomography angiography (CCTA). For the development of effective prevention strategies beyond known risk factors, it is essential to understand the metabolic processes underlying both early and advanced stages of atherosclerosis progression. Plasma metabolomics, which enables comprehensive profiling of small molecules in plasma, offers a promising approach to identify metabolic markers that reflect or influence these processes across the atherosclerosis continuum. Mass spectrometry (MS)-based metabolomics provides high sensitivity and broad coverage, particularly for low-abundance and small metabolites, making it a powerful tool for investigating atherosclerosis pathophysiology. However, few large-scale population studies have examined associations between MS-based metabolomics and atherosclerosis progression. These studies involved limited sample sizes (n<2,500)^2–5^ and lacked detail on the atherosclerotic burden and its progression over time.

We aimed to identify metabolites associated with subclinical coronary atherosclerosis, to evaluate their relevance at later disease stages, and to describe the association of these metabolites with known cardiovascular risk factors. To this end we combined untargeted plasma metabolomics using ultra-performance liquid chromatography–MS with high-quality anatomical assessments of coronary arteries in a large population-based cohort of individuals without prior cardiovascular disease (CVD). We subsequently assessed whether the metabolites linked to subclinical atherosclerosis were also associated with the risk of developing a myocardial infarction within six months, and whether the metabolites were also associated with atherosclerotic burden in patients with a myocardial infarction. Our study design enables a comprehensive assessment of circulating metabolites associated with atherosclerosis across the continuum of disease progression from its earliest detectable stages through a clinically manifested disease and identifies potential markers and biological pathways warranting further investigation.

## Methods

### Data sources

All participants across all cohorts provided written informed consent. The studies were conducted per the Declaration of Helsinki and received approval from the respective ethics committees (Swedish CArdioPulmonary bioImage Study (SCAPIS): Swedish Ethical Review Authority, Dnr 2022-06137-01; Markers of Imminent Myocardial Infarction (MIMI): Regional Ethics Review Board at Uppsala University, Dnr 2016/197; Providing Regional Observations to Study Predictors of Events in the Coronary Tree II (PROSPECT II): Swedish Ethical Review Authority, Dnr 2020-02553; Prospective Investigation of Obesity, Energy and Metabolism (POEM): Regional Ethics Review Board at Uppsala University, Dnr 2009/057).

#### SCAPIS

The Swedish CArdioPulmonary bioImage Study is a population-based study designed to investigate cardiovascular and respiratory diseases, comprising 30,154 individuals aged 50 to 64 years, recruited from 6 university hospitals across Sweden.^6^ Individuals in SCAPIS underwent extensive imaging, including ultrasound of the carotid arteries, computed tomography (CT), and CCTA of the coronary arteries for individuals without contraindications. In addition, participants underwent blood pressure and anthropometric measurements, completed questionnaires on lifestyle, diet (MiniMeal-Q questionnaire^7^), smoking habits, country of birth, and health history, and provided fasting blood samples. Age and sex were extracted from population registers. Blood samples were analyzed for total cholesterol, high-density lipoprotein (HDL) cholesterol, triglycerides, glucose, high-sensitivity C-reactive protein (hsCRP), and creatinine using standard laboratory methods. Non-HDL cholesterol was calculated as total cholesterol minus HDL cholesterol, and estimated glomerular filtration rate (eGFR; mL/min/1.73 m²) was calculated using the Revised Lund–Malmö Study equation.^8^ Medication use was obtained through linkage to the Swedish Prescribed Drug Register in SCAPIS (prescriptions dispensed within the past year). Detailed information on data collection and management is provided in the Supplementary Methods.

The present study focused on individuals without prior CVD events and with complete coronary artery calcium score (CACS) and metabolomics data. Prior CVD events were defined as myocardial infarction, angina, atrial fibrillation, valvular disease, previous bypass surgery or percutaneous coronary intervention, revascularization of other arterial vessels, and stroke. Individuals with missing data were considered to have prior CVD if multiple imputation procedures classified them as prior CVD cases in at least six out of ten imputations.

Subclinical coronary atherosclerosis in SCAPIS participants was evaluated using CT imaging, with detailed methodologies available in previous publications.^6, 9^ Coronary atherosclerosis was assessed using three measures. CACS was derived from a non-contrast CT scan to quantify calcification in the coronary arteries and calculated using syngo.via calcium scoring software (Volume Wizard; Siemens), summing the area of coronary calcification across the entire coronary tree according to the Agatston method.^10^ Segment involvement score (SIS) was assessed using CCTA with iohexol (350 mg I/mL; GE Healthcare) administered at a dosage of 325 mg/kg body weight. CCTA images were reconstructed and analyzed using syngo.via software, following the 18-segment model established by the Society of Cardiovascular Computed Tomography^11^ but focusing on the 11 clinically most relevant segments (segments 1 through 3, 5 through 7, 9, 11 through 13, and 17). Each coronary segment was visually examined for the presence of plaques, and each plaque was characterized. SIS was calculated by counting the total number of coronary segments in which atherosclerotic plaques were detected. This resulted in an ordinal variable with 12 possible levels.^9^ A binary outcome of ≥50% stenosis in at least one coronary vessel was also created based on the degree of coronary stenosis in each segment.

Presence of carotid atherosclerotic plaques in SCAPIS was assessed following a standardized protocol and the Mannheim consensus.^12^ Two-dimensional grayscale ultrasound images were captured using an Acuson S2000 ultrasound scanner and a 9L4 linear transducer (both Siemens) and carotid plaque were defined as absent, unilateral, or bilateral.

#### MIMI

The Markers of Imminent Myocardial Infarction study is a case-cohort study nested within the BBMRI-LPC (Biobanking and Biomolecular Research Infrastructure—Large Prospective Cohorts) collaboration that incorporated data from six cohorts from nine countries across Europe.^4^ Included cohorts were EpiHealth, Trøndelag Health Study (HUNT), Lifelines, the European Prospective Investigation into Cancer and Nutrition—Cardiovascular Disease (EPIC-CVD), Estonian Biobank study, and Malmö Preventive Project. Participants with available blood and data from cohorts included within the BBMRI-LPC were included, and participants with a history of CVD, or renal failure were excluded. Imminent myocardial infarction cases were defined as participants with a first incident acute myocardial infarction, i.e., hospitalizations or deaths primarily due to myocardial infarction, within six months after baseline, when the blood for metabolomics was collected (International Classification of Diseases (ICD)-10: I21; ICD-9: 410.0–410.6, 410.8). Up to four cohort representatives per imminent myocardial infarction case were selected based on sex, age, and study center, using a stratified case-cohort design. Efforts were made to include only type 1 myocardial infarctions by excluding cases with secondary diagnoses such as anemia, tachyarrhythmias, heart failure, renal failure, chronic obstructive pulmonary disease, sepsis, or hypertensive crises. The final study sample included 420 cases and 1,598 subcohort representatives from a total cohort of 169,053 participants. Measurements of body mass index (BMI), blood pressure, lipid levels, glucose, diabetes status, smoking status, and other clinical and demographic variables were harmonized across its participating cohorts.%

#### PROSPECT II

The Providing Regional Observations to Study Predictors of Events in the Coronary Tree II study, is an investigator-sponsored, multicenter, prospective natural history study conducted at 14 hospitals in Denmark, Norway, and Sweden.^13^ The study enrolled patients of any age who had suffered a myocardial infarction within the past four weeks and had successfully undergone intervention for all flow-limiting culprit lesions. Non-culprit lesions of the initial myocardial infarction were measured using intravascular ultrasound and near-infrared spectroscopy (NIRS). Three outcomes measured at study inclusion were considered in the present analyses: 1) pan-coronary lipid, with the lipid core burden index (LCBI) representing the average lipid deposition in the three coronary arteries; 2) pan-coronary plaque volume, representing the average plaque burden in the coronary arteries; 3) presence of a vulnerable high lipid plaque, defined as a lesion with maximum lipid core burden over any 4-mm segment (maxLCBI4mm) ≥ Upper quartile (324.7). LCBI describes the fraction of pixels, generated by NIRS, with a probability of lipid plaques greater than 0.6 divided by all analyzable pixels within the region of interest. Plaque burden was derived using the formula (vessel area–lumen area)/vessel area. Demographic and clinical data were captured in study-specific case report forms (CRFs) for inclusion/exclusion criteria in a standard registry-trial manner.

#### POEM

The Prospective Investigation of Obesity, Energy and Metabolism study is a population-based study conducted in Uppsala, Sweden, aiming to investigate metabolic and cardiovascular health.^14^ Between 2010 and 2016, 502 individuals were enrolled in the study 1 month after turning 50. All participants completed questionnaires and underwent comprehensive physiological assessments after an overnight fast, including blood sampling via a brachial artery cannula, lipid and glucose measurements, body composition analysis using bioimpedance, and carotid artery ultrasonography using an external B-mode ultrasound imaging (Acuson XP128 with a 10-MHz linear transducer, Mountain View, CA, USA). Two outcomes were derived from these measurements: carotid artery intima-media thickness (IMT) and intima-media echogenicity/gray scale median (IMGSM), a measure of intima-media echogenicity. Of the 502 enrolled participants, 487 participants with metabolomic data, valid carotid ultrasonographic scans, required covariates, and no history of CVD (myocardial infarction and stroke; self-reported) were included in the present analysis. Self-reported country of birth was classified as Sweden (yes/no).

### Metabolomics

Fasting plasma samples from SCAPIS participants were collected during their first visit and stored at –80°C until analysis by Metabolon Inc. (Durham, NC, USA), following protocols previously described.^15, 16^ The samples were part of three different batches: Malmo (n=3,864), Uppsala-1 (n=2,841), and Uppsala-2 (n=1,866). In MIMI, samples were from fasting and non-fasting participants. In POEM, fasting plasma samples were also used, while in PROSPECT II non-fasting samples were measured. Metabolite profiling in SCAPIS, PROSPECT II, MIMI, and POEM was conducted using UPLC-MS/MS under multiple conditions: reverse-phase positive and negative electrospray ionization and hydrophilic interaction chromatography with negative electrospray ionization by Metabolon Inc. (Durham, NC, USA), following protocols previously described.^15, 16^. Details on which condition were used for each metabolite is provided in Supplementary Table 4.

In brief, each sample was precipitated with methanol to remove proteins, divided into fractions for each condition, and then dried. The fractions were reconstituted using a specific solvent for each condition. For the positive ion conditions, water and ethanol, acetonitrile, perfluoropentanoic acid, or formic acid were added. For negative ion conditions, samples were eluted with water, methanol, ammonium bicarbonate, acetonitrile, or ammonium formate. Raw data was extracted and further processed using Metabolońs proprietary hardware and software. Compounds were identified by comparison to over 3,300 purified standards and recurring unknown compounds without a confirmed structure, designated as “X-“ followed by a number, based on retention time/index, mass to charge ratio, chromatographic data, and MS/MS spectra. Metabolites were quantified using area-under-the-curve. A correction to each metabolite in each analytical batch across day-runs was applied to correct for inter-day variation from instruments by dividing the peak measurement areas by the median peak area of each metabolite. Then, missing metabolite values were replaced with the minimal non-missing observed value of that metabolite. Metabolites were classified into metabolic classes, referred to as super pathways and sub pathways. In SCAPIS, we compared the levels of cholesterol, creatinine, and glucose measured at the reference laboratory with the measurement performed by Metabolon which indicated good comparability of the measurements.

Metabolites with values below the detection threshold were imputed to the lowest observed value above the detection threshold. Metabolites detected in fewer than 30 participants per batch, not present in all batches, or annotated as drug metabolites in the sub pathway classification and detected in all participants were excluded from the analysis (k=245). Among the remaining metabolites, those annotated as drug metabolites in the sub pathway classification or with a prevalence below 2% were converted into binary variables (k=40), while all others were log2-transformed (k=1,131) for all analyses.

### Statistical analysis

In SCAPIS, missing values of subclinical atherosclerosis measures, metabolites, covariates, and the primary outcome SIS were imputed using multiple imputation by chained equations (MICE)^17^ as implemented in the mice package in R (version 3.16.0)^18^ based on 8,571 individuals (52% female, mean age 57.5) with available CACS, sex, age, and birth country data, including those with self-reported CVD (n=422). Since some participants had contraindications for the contrast infusion (e.g., impaired kidney function), and hence had no CCTA and SIS recording, we decided to expand the dataset based on available CACS measurements by imputations, including eGFR in the imputation model.^19^ SIS was available in 7,269 individuals before imputation. Proportional odds regression was used for imputing SIS, while other variables were imputed using predictive mean matching with 5 nearest neighbours. Imputations were performed separately for all three batches (Malmö, Uppsala-1, Uppsala-2), with 10 imputations being performed in total for each batch. Rubin’s rules were used to calculate pooled standard errors for all analyses on imputed data of each batch. Batch-specific estimates were combined using an inverse-variance weighted fixed-effect meta-analysis.

#### Association between plasma metabolites and subclinical coronary atherosclerosis in SCAPIS

Multivariable regression models were used to assess the association between plasma metabolites and subclinical atherosclerosis in SCAPIS, using R version 4.3.1. Each metabolite was analyzed individually as the independent variable, with three different measurements of subclinical atherosclerosis as the dependent variables. Ordinal regression was used to examine the association with SIS, an ordinal variable with 12 levels. Linear regression was applied to evaluate the association between plasma metabolites and CACS. To correct for skewness, CACS was transformed using the natural logarithm after adding a pseudo-count (CACS+1). Logistic regression with Firth correction was applied to assess the association with stenosis ≥50%, a binary outcome. Abundant metabolites were log2-transformed, while metabolites with low prevalence (i.e., <2%) or representing drug compounds were categorized as binary (present/absent).

Three models were evaluated. Model 1 included adjustments for age, sex, and country of birth (i.e., non-modifiable risk factors). Model 2 further adjusted for smoking status (3 categories), passive smoking exposure, BMI, waist-to-hip ratio, eGFR, systolic blood pressure, diastolic blood pressure, and antihypertensive medication. To obtain metabolites that are independent of clinical measures of lipidemia, glycemia and inflammation as well as to better control for smoking, we also examined Model 3 which included additional adjustments for cotinine metabolite levels, HDL cholesterol, non-HDL cholesterol, hsCRP, glucose, and medications for diabetes and dyslipidemia.

Multiple testing was controlled using the Benjamini-Hochberg FDR set at 5% (q-value<0.05) for each outcome separately and for all tested metabolites (k=1,171).^20^ We applied the threshold of q-value<0.05 for all further multiple testing corrections. Plasma metabolites associated with SIS at a q-value<0.05 were selected for further exploration with the other outcomes across the atherosclerosis continuum.

Sensitivity analyses were performed to test the robustness of the associations by comparing results from the main analysis (SIS), to results when excluding current and former smokers, and comparing results from the main analysis based on imputed data with complete case analysis.

To assess the influence of adding each covariate, we performed additional modeling by introducing one variable at a time for metabolites associated with SIS at a q-value < 0.05, following either alphabetical or reverse alphabetical order.

#### Metabolomic class enrichment analysis

Enrichment analysis of the SIS results in SCAPIS was conducted at the metabolite subpathway level using enrichment analysis with the fgsea R package (version 1.26.0)^21^ on ranked p-values, incorporating all associations. Separate analyses were performed for positive and negative regression coefficients.

#### Association between atherosclerosis-related metabolites and imminent myocardial infarction

Weighted, stratified Cox proportional-hazards models were used to assess the association between coronary atherosclerosis-associated metabolites and incident events of myocardial infarction in MIMI. Inverse sampling probability weights (Borgan II) accounted for the case–cohort design within a stratified model, allowing for different baseline hazard shapes across the six MIMI cohorts. A robust variance estimator (Huber–White) was applied to ensure reliable standard errors. Associations were adjusted for age, sex, country of residence, smoking status, log2-transformed cotinine, waist circumference, log2-transformed creatinine, and prevalent diabetes. These variables were chosen to mimic the variables in the SCAPIS analysis. Information on passive smoking exposure, antihypertensive medication use, dyslipidemia medication use, and hsCRP was unavailable in MIMI and could therefore not be included as covariates. Statistical significance is reported as nominal (p-value<0.05) and after correcting for multiple comparisons by using the Benjamini-Hochberg method with an FDR cut-off of 5%.

#### Association between atherosclerosis-related metabolites and atherosclerosis burden in myocardial infarction patients

Furthermore, associations of the 12 coronary atherosclerosis-associated metabolites with three measures of atherosclerosis burden in PROSPECT II were investigated. Associations with two continuous measures: 1) pan-coronary lipid, 2) pan-coronary plaque volume were estimated using multivariable linear regressions, and 3) a binary measure of presence of high-risk vulnerable plaques with logistic regressions. Associations were adjusted for age, sex, country of birth, smoking status, BMI, medication for hyperlipidemia, HDL cholesterol, non-HDL cholesterol, hsCRP, and plasma glucose. Information on passive smoking exposure, cotinine metabolite levels, waist-to-hip ratio, eGFR, systolic and diastolic blood pressure, antihypertensive medication use, and diabetes medication use was unavailable in PROSPECT II and could therefore not be included as covariates. Statistical significance is reported as nominal and after correcting for multiple comparisons by using the Benjamini-Hochberg method with an FDR cut-off of 5% applied for each outcome separately.

#### Association of atherosclerosis-related metabolites with determinants of CVD risk and carotid phenotypes

Partial Spearman correlations were calculated to examine whether the coronary atherosclerosis-associated metabolites were correlated with cardiovascular risk factors, medications, and carotid phenotypes, adjusting for age, sex, and country of birth. For carotid phenotypes, correlations with carotid plaque were performed using SCAPIS data, and correlations with IMT and IMGSM in POEM. Rank-based inverse normal transformation was applied to IMT and IMGSM. In SCAPIS, Spearman’s correlations were estimated per batch, with pooling performed using Rubin’s rules after Fisher’s z-transformation of the correlation coefficients and subsequently meta-analyzed using inverse-variance weighted fixed-effect meta-analysis. For POEM, Spearman’s correlations were estimated using the entire dataset. Statistical significance is reported as nominal (p-value<0.05) and after correcting for multiple comparisons by using the Benjamini-Hochberg method with an FDR cut-off of 5% applied for each outcome separately.

## Results

The overall study design is depicted in Figure 1. In brief, we first investigated the associations between 1,171 circulating metabolites and atherosclerosis in 8,146 individuals free of clinical manifestations of CVD. Secondly, we assessed whether the associated metabolites were also associated with (i) the risk of imminent myocardial infarction (420 events) in a case-cohort study, defined as a first incident acute myocardial infarction within six months after blood collection for metabolomics, and (ii) non-culprit atherosclerotic burden in a separate, independent cohort of 898 patients with previous myocardial infarction. Lastly, we describe the association of these metabolites with cardiovascular risk factors, including carotid phenotypes.

**Figure 1.**
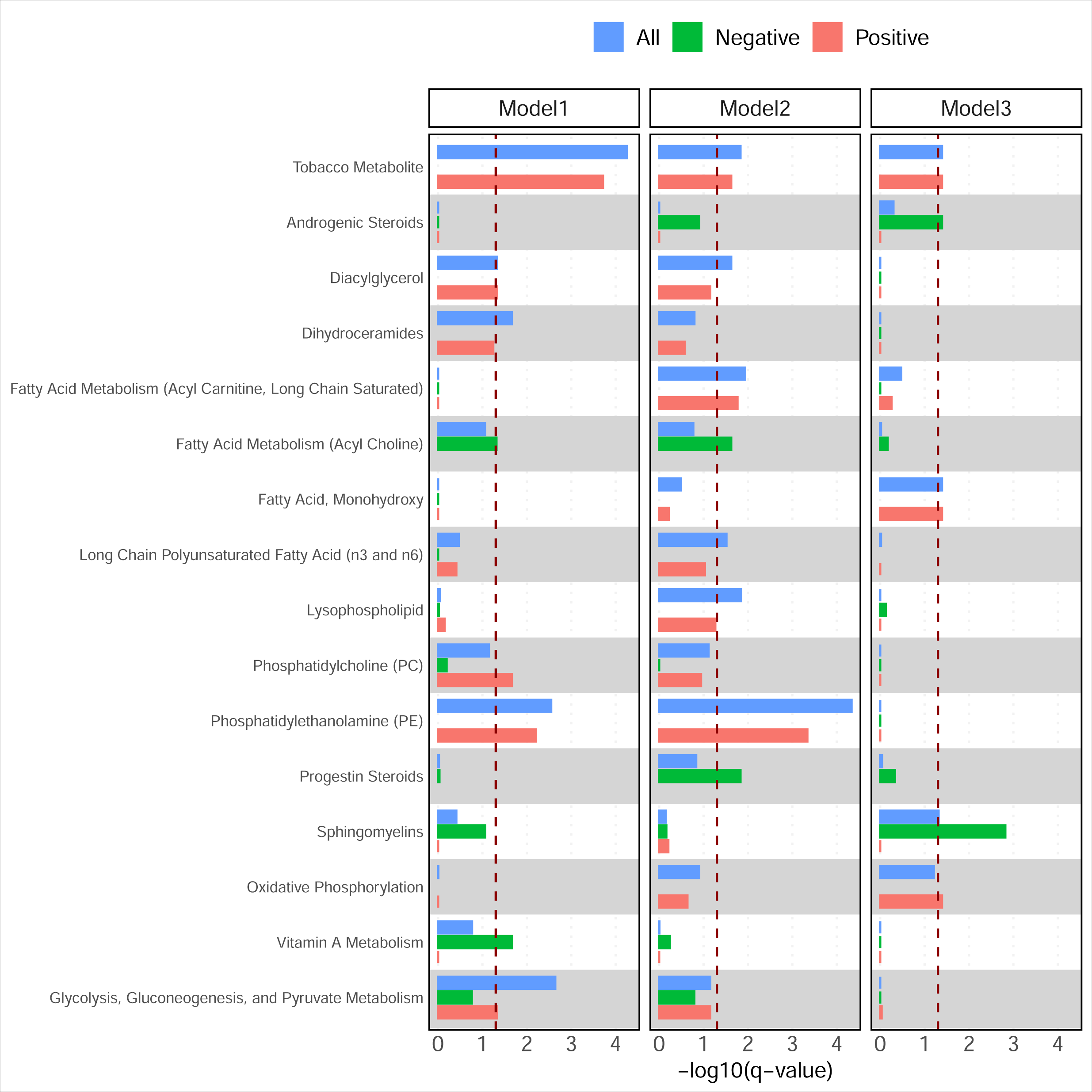
Study design and number of participants in each analysis. CACS: coronary artery calcium score; CT: computed tomography; CVD: cardiovascular disease; MIMI: Markers of Imminent Myocardial Infarction; PROSPECT II: Providing Regional Observations to Study Predictors of Events in the Coronary Tree II; SCAPIS: Swedish CArdioPulmonary bioImage Study; SIS: segment involvement score.

### Twelve plasma metabolites are robustly associated with subclinical coronary atherosclerosis

We investigated the associations of 1,171 circulating metabolites with CCTA-based SIS in 8,146 SCAPIS participants without prior CVD (53.1% female, mean age 57.4 years; Table 1), of whom 42.6% had at least one coronary atherosclerosis lesion at baseline. SIS represents the number of coronary tree segments affected by atherosclerosis. We chose this measure over CACS score and presence of stenosis because SIS captures the overall burden and distribution of both calcified and non-calcified plaque across the entire coronary tree, providing a more comprehensive measure of atherosclerosis disease. We specified three sets of covariates for the analysis in SCAPIS. The choice to include three models was to address challenges in defining a unified covariate adjustment suitable for all types of metabolites and because we wanted to understand how the addition of different types of covariates affected the model. Model 1 was a minimal model adjusted for age, sex, and country of birth. After multiple imputation and corrections for multiple testing (false discovery rate, FDR, of 5%), we identified 524 associated metabolites (Supplementary Table 1). In Model 2, we additionally adjusted for smoking, anthropometric traits, eGFR, blood pressure, and antihypertensive medication, which reduced the number of associated metabolites to 167 (Supplementary Table 1). The metabolites with the strongest associations in Model 2 were predominantly lipids within the ceramide and phospholipid subclasses, as also shown by formal enrichment analyses, which additionally identified enrichment for tobacco metabolite pathways (Figure 2). We subsequently applied Model 3, additionally adjusting for lipids, glycemic markers, lipid-lowering and antidiabetic medications, hsCRP, and cotinine, a biomarker of smoking (Supplementary Table 1). Using this model, we identified 12 associated metabolites, of which 8 showed positive and 4 negative associations. These associations included phosphate, malate, six lipids independent of cholesterol levels (three of which were sphingomyelins), three amino acids, and an uncharacterized metabolite X-25790, which Metabolon Inc. (Durham, NC, USA) considers possibly a nucleoside derivative (Figure 3, Supplementary Figure 1). Its observed [M+H]^+^ ion at m/z 247.0923 (<1 ppm from theoretical) and isotopic pattern indicate the molecular formula C9H14N2O6, identical to that of 5,6-dihydrouridine. However, analysis of an authentic 5,6-dihydrouridine standard excluded this identity.

**Figure 2.**
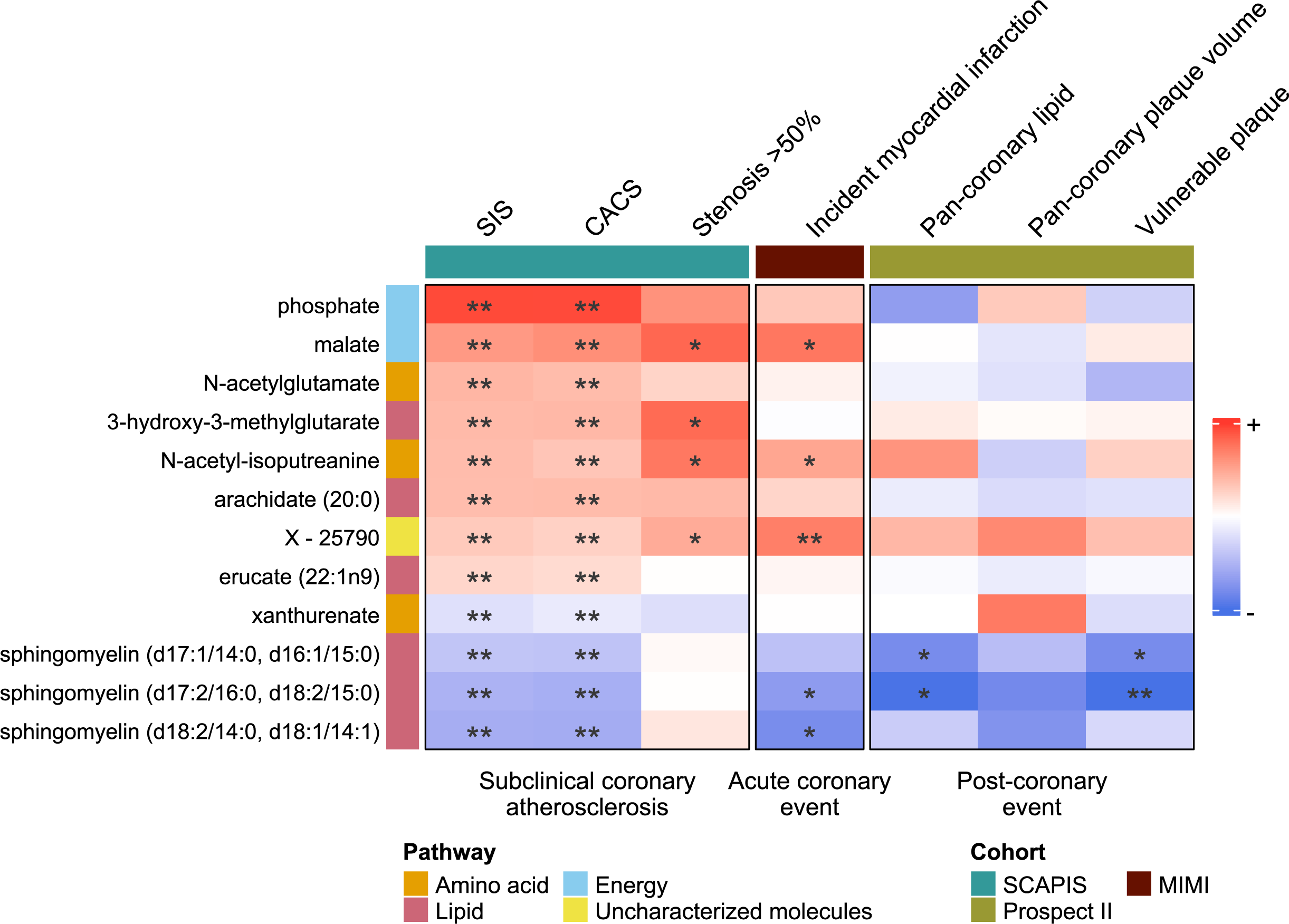
Enrichment analysis of metabolites associated with subclinical atherosclerosis, measured by segment involvement score (SIS), across Model 1-3. Bars extending beyond the dashed line indicate significant associations (q-value<0.05). Blue bars represent enrichment analysis using all p-values from the associations between metabolites and SIS. Green bars correspond to enrichment analysis using p-values from associations with negative coefficients, while red bars correspond to those with positive coefficients. Model 1 was adjusted for age, sex, and country of birth. Model 2 included additional adjustments for smoking status, anthropometric traits, estimated glomerular filtration rate (eGFR), blood pressure, and use of antihypertensive medication. Model 3 was further adjusted for lipid levels, glycemic markers, use of lipid-lowering and antidiabetic medications, C-reactive protein, and cotinine.

**Figure 3.**
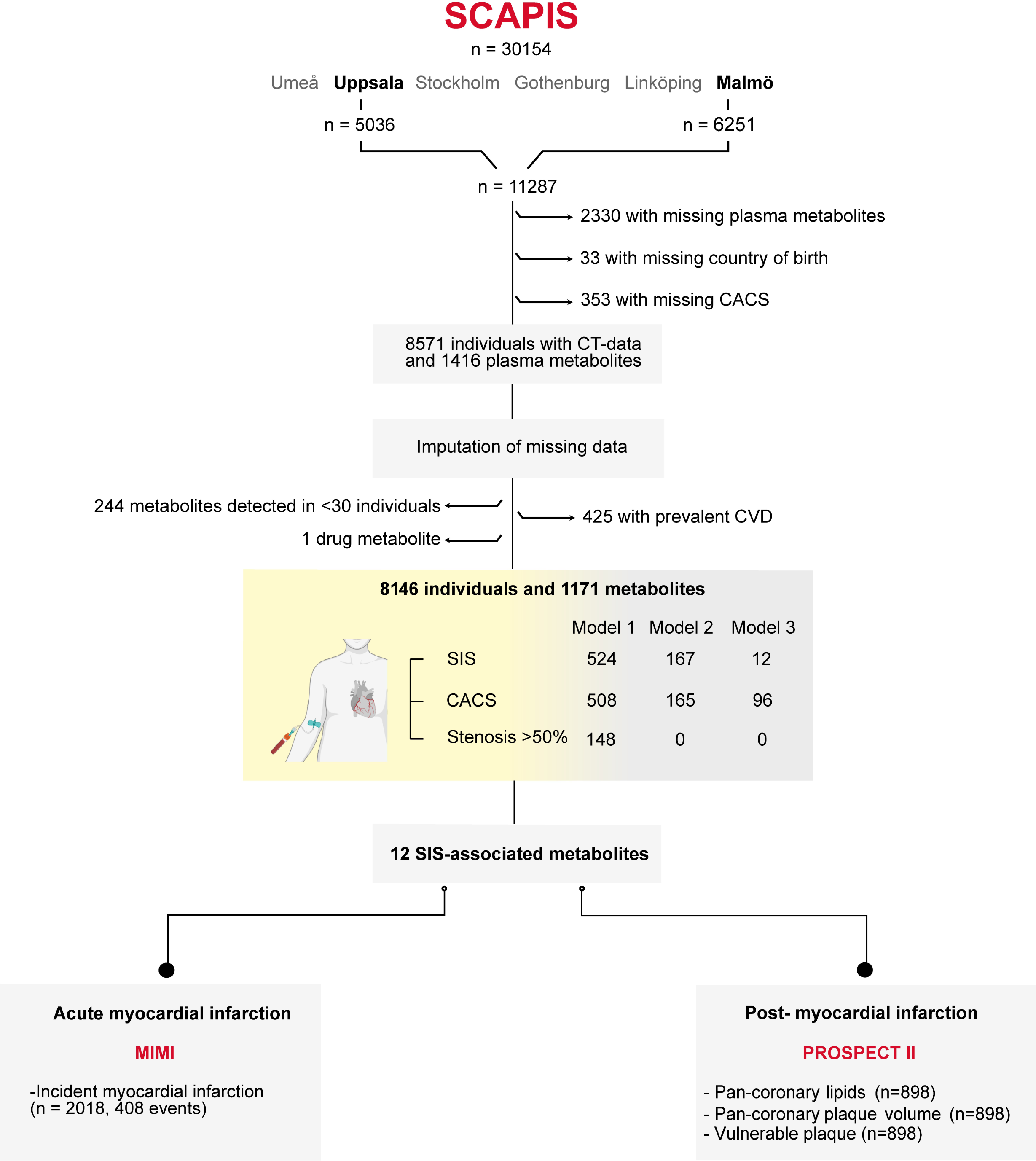
Association of metabolites with atherosclerosis outcomes in fully adjusted analyses. For visualization purposes, beta coefficients for the outcomes (shown as columns) were scaled (i.e., SD-transformed); color intensity reflects the magnitude of the scaled beta coefficient for a metabolite compared to the other metabolites for the same outcome. For segment involvement score (SIS), stenosis >50%, carotid plaque, and vulnerable plaque, beta coefficients were derived from log-transformed odds ratios. For incident myocardial infarction, beta coefficients were derived from log-transformed hazard ratios. Models in SCAPIS were adjusted for age, sex, and country of birth, smoking status (3 categories), passive smoking exposure, body mass index (BMI), waist-to-hip ratio, estimated glomerular filtration rate (eGFR), systolic blood pressure, diastolic blood pressure, and antihypertensive medication, cotinine metabolite levels, high-density lipoprotein (HDL) cholesterol, non-HDL cholesterol, C-reactive protein, glucose, and medications for diabetes and dyslipidemia. Models in MIMI were adjusted for country of residence, smoking status (never, former, current), log2-transformed cotinine, waist circumference, log2-transformed creatinine, and prevalent diabetes. Models in PROSPECT II were adjusted for age, sex, country of birth, smoking status, BMI, medication for hyperlipidemia, HDL cholesterol, non-HDL cholesterol, C-reactive protein, and plasma glucose. Statistical significance: **false discovery rate (FDR) q < 0.05; *nominal p < 0.05 but FDR-adjusted q ≥ 0.05; no stars: p ≥ 0.05. CACS: coronary artery calcium score; MIMI: Markers of Imminent Myocardial Infarction; PROSPECT II (P-II): Providing Regional Observations to Study Predictors of Events in the Coronary Tree II; SCAPIS: Swedish CArdioPulmonary bioImage Study; SIS: segment involvement score.

**Table 1.** Description of the included studies.

|  | SCAPIS<br>(n=8,146) | MIMI (n=2,018) |  | PROSPECT II<br>(n=898) | POEM (n=487) |
| --- | --- | --- | --- | --- | --- |
|  |  | Cases<br>(n=420) | Subcohort<br>(n=1,598) |  |  |
| Male (%) | 46.9 | 63.8 | 63.8 | 49.9 | 82.8 |
| Age, y | 57.4 (4.3) | 64.1 (12.1) | 63.0 (12.0) | 50.0 (0) | 63.0 (55.0-70.0) |
| Current smoking (%) | 12.8 | 32.7 | 24.4 | 9.5 | 31.8 |
| Body mass index, kg/m<sup>2</sup> | 27.0 (4.4) | 27.6 (4.2) | 27.4 (4.1) | 26.4 (4.2) | 27.1 (24.8-30.2) |
| Waist-to-hip ratio | 0.92 (0.09) | NA | NA | 0.90 (0.07) | NA |
| HDL cholesterol, mmol/L | 1.57 (0.48) | 1.3 (0.4) | 1.3 (0.4) | 1.40 (0.39) | 1.1 (1.0-1.4) |
| Non-HDL cholesterol,<br>mmol/L | 4.04 (1.06) | 4.7 (1.3) | 4.5 (1.1) | 3.98 (0.98) | 3.9 (3.1-4.7) |
| Systolic blood pressure,<br>mmHg | 124 (16.1) | 142.1<br>(21.1) | 139.7 (19.8) | 126 (16.5) | NA |
| Diastolic blood pressure,<br>mmHg | 75.9 (9.8) | 80.4 (12.7) | 79.2 (11.7) | 77.0 (10.2) | NA |
| Plasma glucose, mmol/L | 5.7 (5.3-6.1) | 6.0 (2.0) | 5.8 (1.9) | 4.5 (4.2-4.8) | 6.1 (5.4-7.05) |
| C-reactive protein, mg/L | 1.1 (0.6-2.3) | NA | NA | NA | 3.4 (1.5-6.1) |
| Estimated GFR,<br>mL/min/1.73 m<sup>2</sup> | 78.2 (9.54) | NA | NA | 83.9 (9.65) | NA |
| Serum creatinine, µmol/L | 75.0 (66.0-85.0) | NA | NA | 75.0 (65.0-84.0) | 78.7 (69.0-90.2) |
| Prevalent diabetes (%) | 7.8 | 12.6 | 9.0 | 1.6 | 12.1 |
| Antihypertensives (%) | 21.9 | NA | NA | 7.8 | 27.3 |
| Lipid medication (%) | 8.9 | NA | NA | 3.7 | 24.9 |
| Diabetes medication (%) | 3.8 | NA | NA | 1.2 | 10.5 |

Values are presented as mean (standard deviation), median (25<sup>th</sup>-75<sup>th</sup> percentile), or percentage. GFR: glomerular filtration rate; HDL: high density lipoprotein; MIMI: Markers of Imminent Myocardial Infarction; NA: not available; POEM: Prospective Investigation of Obesity, Energy and Metabolism; PROSPECT II: Providing Regional Observations to Study Predictors of Events in the Coronary Tree II; SCAPIS: Swedish CARDioPulmonary bioImage Study.

Notably, the inverse associations of sphingomyelins with SIS were strengthened in Model 3 compared to Model 2, also supported by the enrichment analysis (Figure 2). Further modelling showed that this was mainly due to the addition of the covariate non-HDL (Supplementary Figure 2). Sensitivity analyses of the 12 metabolites, restricted to the never smokers, yielded consistent results, supporting the robustness of the associations (Supplementary Figure 3a). Additional analysis without imputation in 6,489 participants with complete data in metabolites, imaging and covariates yielded consistent results (Supplementary Figure 3b).

We further investigated the association of all metabolites with CACS, and ≥50% stenosis in at least one coronary vessel as secondary outcomes. Associations between metabolites and CACS and enrichments closely mirrored those observed for SIS, but with a larger number of associated metabolites (Supplementary Table 2). For the binary outcome of ≥50% stenosis (6% of the population), we identified fewer associated metabolites; however, their estimates were largely correlated with SIS estimates, albeit only weakly for Model 3 (Supplementary Table 3, Supplementary Figure 4a-f).

In summary, our analysis provides robust evidence for the association of 12 metabolites, spanning 4 broad metabolite classes, with subclinical coronary atherosclerosis after extensive adjustment for cardiovascular risk factors and medication use, hereinafter referred to as coronary atherosclerosis-associated metabolites.

### Malate, N-acetyl-isoputreanine, X-25790 and two sphingomyelins are associated with incident myocardial infarction

In the MIMI stratified case-cohort study, which included 2,018 participants without prior CVD at baseline across 6 studies, and 420 myocardial infarction events occurring within the first 6 months of follow-up, we assessed whether coronary atherosclerosis-associated metabolites were associated with an increased risk of myocardial infarction within 6 months. Individuals with events had a similar age, body mass index, and proportion of males compared to those without events; however, 12.6% of those with events had prevalent diabetes, while in those without events, this was 9.0%. We applied a similar covariate adjustment set to Model 3. Malate, N-acetyl-isoputreanine, and X-25790 were positively associated, while two sphingomyelins were negatively associated with the risk of imminent myocardial infarction (p-value<0.05) (Figure 3). All these associations were consistent in their direction compared to the associations in SCAPIS with subclinical atherosclerosis.

### Sphingomyelins are inversely associated with high-risk plaques

We further examined the associations of the coronary atherosclerosis-associated metabolites with plaque characteristics in the PROSPECT II study (n=898) using a similar set of covariates as in Model 3. The PROSPECT II study enrolled patients of any age who had suffered a myocardial infarction within the past 4 weeks and had successfully undergone percutaneous coronary intervention at 14 hospitals in Denmark, Norway, and Sweden. Non-culprit lesions of the initial myocardial infarction were measured using intravascular ultrasound and NIRS. In PROSPECT II, two of the sphingomyelins, sphingomyelin (d17:1/14:0, d16:1/15:0) and sphingomyelin (d17:2/16:0, d18:2/15:0), were inversely associated with the pan-coronary lipid measured as LCBI and presence of vulnerable high-lipid plaque (Figure 3). These results indicate that sphingomyelins are not only inversely associated with the subclinical coronary atherosclerosis burden in those without CVD but also inversely associated with coronary artery atherosclerosis burden and high-risk plaques in myocardial infarction survivors.

### Atherosclerosis-associated metabolites are correlated with cardiovascular risk factors

To explore correlations of the coronary atherosclerosis-associated metabolites with carotid phenotypes, cardiovascular risk factors and dietary factors, we calculated Spearman partial correlation adjusted for age, sex, and birth country in SCAPIS, similar to Model 1. We observed multiple associations including a strong negative correlation of the uncharacterized metabolite X-25790 with eGFR, and positive correlations between the three sphingomyelins with HDL and non-HDL cholesterol as well as negative correlations with lipid-lowering medication (Supplementary Figure 5). Three carotid phenotypes were examined: ultrasound-derived carotid plaque (defined as the number of carotid arteries with >0 plaque with focal intima-media thickness >1.5 mm) in SCAPIS, as well as with carotid IMT and IMGSM in the POEM study of 487 CVD-free individuals. In SCAPIS, phosphate was positively correlated with carotid plaque, while malate was negatively correlated (Supplementary Figure 5). In POEM, two of the sphingomyelins were inversely associated with IMGSM.

## Discussion

In this extensive multicohort study integrating untargeted plasma metabolomics with detailed imaging across the atherosclerosis continuum, we identified a set of 12 metabolites, phosphate, malate, 6 lipids, 3 amino acids, and 1 uncharacterized metabolite, robustly associated with subclinical coronary atherosclerosis after extensive adjustment for traditional cardiovascular risk factors, lipid and glycemic markers, and an objective biomarker of smoking (cotinine), supporting the role of specific metabolic alterations beyond established risk pathways. Several of these metabolites were also associated in independent cohorts with the risk of myocardial infarction within 6 months after blood sampling as well as with plaque burden and presence of high-risk vulnerable plaque.

Among the key findings, sphingomyelins emerged as metabolites inversely associated with coronary atherosclerosis across both subclinical (SCAPIS) and clinical (PROSPECT II) settings, including plaque volume and lipid content of the coronary tree. Furthermore, these sphingomyelins were also inversely associated with a lower risk of myocardial infarction in MIMI. We observed that the inverse associations of sphingomyelins were more prominent after adjustment for non-HDL cholesterol. A potential mechanism involves apolipoprotein C-III (apoC-III), an established cardiovascular risk factor.^22, 23^ LDL particles enriched in apoC-III contain lower amounts of sphingomyelin and are more susceptible to sphingomyelinase-mediated hydrolysis, aggregation, and arterial retention. Reduced circulating sphingomyelin levels may therefore serve as a marker of apoC-III-rich, pro-atherogenic LDL particles, consistent with the strengthening of the inverse association after non-HDL-cholesterol adjustment observed in our study.^22, 24, 25^ We did not measure apoC-III in this study, and this explanation remains speculative.

Previous studies have shown divergent results, which we speculate could be partly due to differences in model specifications or due to heterogeneity across sphingomyelin species. Plasma levels of sphingomyelins have been positively associated with prevalent coronary artery disease^26, 27^ and subclinical atherosclerosis,^28, 29^ but others have also reported negative associations with incident CVD^3, 30–32^ or markers of subclinical CVD.^33^ Several studies have also suggested that distinct molecular species of sphingomyelins are differently associated with CVD, with sphingomyelins species containing highly polyunsaturated and long-chain saturated fatty acids (e.g. 18:2) being associated with a lower risk of CVD, and sphingomyelin species with palmitic acid (16:0) being associated with higher risk.^34–37^ In this study, we mainly observed inverse associations in species containing d18:2 rather than d18:1. However, in our study, for most of the sphingomyelin molecules the exact isoform could not be determined. Sphingomyelins are important constituents of the membranes of lipoprotein particles, with ApoB-containing lipoproteins (e.g., low-density lipoproteins (LDL) cholesterol) containing the highest concentrations of sphingomyelins.^38^ The metabolomics measurement of plasma sphingomyelins therefore renders a high correlation of plasma sphingomyelins with circulating lipoprotein concentrations. Our non-HDL-cholesterol-adjusted estimates likely reflect the association between the relative content of sphingomyelins in pro-atherogenic lipoproteins and cardiovascular outcomes. Taken together, these results suggest that a high abundance of specific sphingomyelins in non-HDL cholesterol species may reflect differences in lipoprotein sphingomyelin composition. While the underlying mechanisms remain to be elucidated, these findings generate the hypothesis that sphingomyelin content in ApoB-containing lipoproteins may be related to atherosclerotic burden. Sphingomyelin content in lipoprotein membranes may affect cholesterol synthesis and uptake^39^, and can also impact the fluidity of the lipoprotein membrane.

Several other metabolites were directly associated with coronary atherosclerosis in the present study. Our finding of a positive association between malate and all three subclinical atherosclerosis as well as incident myocardial infarction is consistent with a possible role of the tricarboxylic acid (TCA) cycle intermediates in CVD pathophysiology. Malate is one of the intermediate metabolites involved in the tricarboxylic acid cycle (TCA, also called Krebs cycle), a central pathway in energy production. Circulating malate is also provided by malate-aspartate shuttle, particularly active in the hepatic metabolism.^40^ Some studies have shown that increased plasma levels of TCA cycle intermediates, are positively associated with prevalent CVD and CVD risk factors,^41–43^ with one study linking dysregulated TCA cycle to cardiovascular events.^5^ In the PREDIMED study, higher baseline levels of malate were associated with incident heart failure and atrial fibrillation,^44^ non-fatal stroke and myocardial infarction.^45^ Other studies using plasma and urine samples reinforce this association, linking malate to atherosclerosis and CVD risk.^46, 47^

One of the strongest and most consistent associations observed across cohorts in the present study was for the metabolite X-25790. Although the compound’s exact identity remains unconfirmed, current evidence most strongly supports its annotation as a nucleoside-like molecule with the molecular formula C₉H₁₄N₂O₆. The metabolite X-25790 was positively correlated with acute coronary events when compared to healthy controls in a retrospective analysis from a case-control study,^48^ corroborating the findings of our study. Furthermore, our study also observed a strong negative correlation between X-25790 and eGFR, suggesting a link to kidney function.

In this study, N-acetyl-isoputreanine was associated with subclinical coronary atherosclerosis and an increased risk of myocardial infarction. N-acetyl-isoputreanine and its related metabolite, N1-acetylisoputreanine-γ-lactam, have been described as a plausible end-product of polyamine metabolism.^49^ N1-acetylisoputreanine-γ-lactam is formed through the oxidation of N1-acetylspermidine by copper-dependent amino oxidases, followed by aldehyde dehydrogenase (ALDH) enzyme activity.^49^ An analogous pathway has been proposed for the N-acetyl-isoputreanine, although this has not been experimentally confirmed.^49^ The specific ALDH isoform involved in this pathway has not been described, but ALDH2 has been extensively linked to atherosclerosis. ALDH2 enzyme is involved in the detoxification of alcohol-derived acetaldehyde and endogenous aldehydes. Reduced ALDH2 enzyme activity has been associated with higher risk of coronary artery disease, myocardial infarction, stroke, and peripheral artery disease, as well as altered cholesterol synthesis, foam cell formation, lower circulating HDL-cholesterol and reduced plaque stability.^50^

Phosphate showed positive associations with atherosclerosis and are in line with results from a recent meta-analysis of observational studies showing positive associations of serum phosphate levels with cardiovascular disease risk.^51^ In the Rotterdam Study, also included in the meta-analysis, higher serum phosphate levels are associated with increased CACS in the general population, with more pronounced effects observed in men.^41^ A Mendelian randomization analysis conducted in the same study further supported a causal relationship between serum phosphate and CACS, even among individuals without hyperphosphatemia, chronic kidney disease, or existing CVD.^52^

This study represents one of the largest efforts to date integrating untargeted plasma metabolomics with imaging-derived measures of subclinical and clinical atherosclerosis, across studies and along different stages of the atherosclerosis continuum. The use of high-resolution coronary CT angiography and coronary calcium scoring in over 8,000 individuals from the SCAPIS cohort, along with assessment of atherosclerotic traits in three independent cohorts, enhances the robustness and generalizability of the findings. We used full cohort data of SCAPIS to identify the metabolites associated with atherosclerosis; this approach has been shown to be more powerful than the discovery/replication cohort approach.^53^ Adjustment for a comprehensive set of confounders, including objective markers of smoking and lipid profiles, further strengthens the validity of the associations. Replication across diverse atherosclerosis-related phenotypes, from subclinical coronary plaque to imminent myocardial infarction, supports the relevance of the identified metabolites across the disease continuum. Nonetheless, several limitations merit consideration. First, the outcome assessed across cohorts capture subclinical atherosclerosis phenotypes, acute cardiovascular events and non-culprit atherosclerotic burden, which may not fully share the same underlying metabolomic mechanisms. Second, many metabolite features, including one of our main findings, remain incompletely characterized or uncharacterized, which limits biological interpretation. Moreover, this study used untargeted metabolomics based on methanol-based protein precipitation which may result in incomplete recovery of hydrophobic lipid species, including sphingomyelins, compared with other extraction methods specifically optimized for lipidomics. Our most exhaustive model was extensively adjusted for known risk factors; this approach might be too conservative and as a result only few associated metabolites remained to be associated with subclinical atherosclerosis. However, our approach also helps identify select metabolites for future studies to prioritize. Third, the observational design precludes strong causal inference, and residual confounding, particularly related to smoking exposure, cannot be fully excluded. Using Mendelian randomization for causal inference is not appropriate for most metabolites due to the presence of weak genetic instruments and/or high pleiotropy. Fourth, heterogeneity in fasting status and sample handling across cohorts might have introduced variability and resulted in loss of statistical power; this might explain few metabolites not associated with the atherosclerotic burden in PROSPECT II, although these metabolites might have been impacted by disease event or treatment in individuals with recent myocardial infarction. Others have shown that metabolomic profiles are relatively stable over time and that fasting status has a small impact.^54^ Nevertheless, in the present study consistent directions of effect were observed across cohorts particularly for the sphingomyelins. Fifth, participants in the studies were largely European ancestry, and replication in other ancestries is needed. Finally, the analysis of stenosis was likely underpowered due to the low prevalence of significant stenosis in this population. Future research should explore the causal role of these metabolites and assess their potential as biomarkers in risk prediction models or therapeutic targeting.

Our results highlight the potential of plasma metabolomics in identifying markers that reflect atherosclerotic disease. Future work should evaluate the mechanistic role of these metabolites, which could lead to new targets for the prevention or treatment of atherosclerosis.

## Acknowledgments

The computations and data handling were enabled by resources in project sens2019512 and sens2020005 provided by the National Academic Infrastructure for Supercomputing in Sweden (NAISS) at Uppsala Multidisciplinary Center for Advanced Computational Science (UPPMAX), funded by the Swedish Research Council through grant agreement no. 2022-06725.

## Data availability statement

The pseudonymized SCAPIS metabolite and phenotype data supporting the conclusions of this article were provided by the SCAPIS central data office and are not shared publicly due to confidentiality and ethical restrictions. Data will be shared only after permission from the Swedish Ethical Review Authority (https://etikprovningsmyndigheten.se) and from the data access board (https://www.scapis.org/data-access). Similarly, access to pseudonymized metabolite and phenotype data from the MIMI study, PROSPECT II, and POEM requires approval from the Swedish Ethical Review Board and the respective data access boards.

Publicly available software was used for the analyses described in the Methods section. The source code and summary data supporting all figures and analyses are available on Github at: https://github.com/MolEpicUU/metab_athero.

