## Supplementary Material for "Multicohort assessment of plasma metabolomic measurements across the atherosclerosis continuum"

### SUPPLEMENTARY METHODS

#### Details on data collection in SCAPIS

Smoking status was determined through a survey asking participants whether they smoked, with response options: "never," "regular," "ex-smoker," "occasional," or "unwilling/unable to reply." For analysis, regular and occasional smokers were classified as current smokers. Passive smoking exposure was assessed based on a separate question about cohabitants' smoking behavior, categorized by duration ("no", "<10 years", "10–20 years", ">20 years").

Participants were grouped into four categories: Scandinavia (Sweden, Denmark, Norway, Finland), non-Scandinavian Europe, Asia, and Other.

Systolic and diastolic blood pressure were measured in both arms using an automatic device (Omron M10-IT, Omron Healthcare, Kyoto, Japan) after five minutes of rest in the supine position. A second measurement was taken after at least one minute, and the final values were averaged from the arm with the highest mean systolic blood pressure.

Data on medications for hypertension, lipid-lowering medication, and glucose-lowering medication were obtained from the Swedish Prescribed Drug Register, covering prescriptions issued in the year before baseline. Medications were classified using Anatomical Therapeutic Chemical (ATC) codes. Antihypertensive medications included ATC codes C02 (antihypertensive drugs), C03A and C03EA01 (thiazide diuretics), C07 (beta-blockers), C08C (selective calcium antagonists with mainly vascular effects), and C09 (agents acting on the renin-angiotensin system). Lipid-lowering medications included statins (C10AA), fibrates (C10AB), bile acid sequestrants (C10AC), and other lipid-modifying agents (C10AX). Diabetes medications included insulin and analogs (A10A) or other glucose-lowering drugs excluding insulin (A10B).

Dietary intake was assessed using the MiniMeal-Q food frequency questionnaire,[1] which captures information on various food items. Carbohydrate and protein intake were energy-adjusted as a percentage of total energy intake. Participants with implausible energy intake values – < 500 or >5000 kcal/day for women and < 550 or >6000 kcal/day for men – were classified as misreporters, and their dietary data were set as missing.

### SUPPLEMENTARY FIGURES

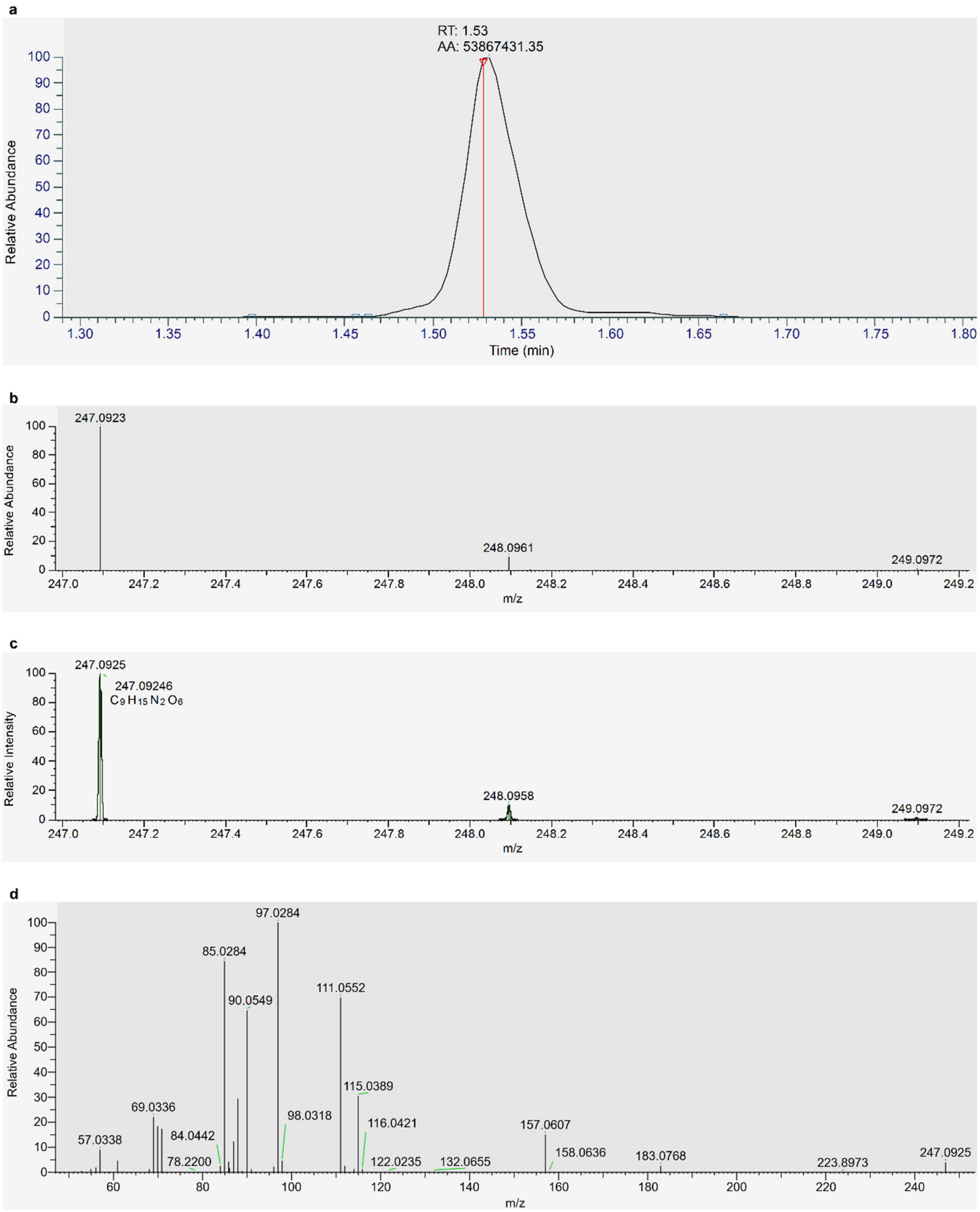


**Supplementary Figure 1.** Mass spectrometric characterization of the uncharacterized metabolite X-25790 (observed m/z 247.0925). (a) Extracted ion chromatogram for m/z 247.0925 using a ±5 ppm mass tolerance. The red vertical line indicates the retention time at the chromatographic peak apex (RT = 1.53 min), which was used for spectral extraction. (b) Experimental MS1 isotopic distribution of the precursor ion at m/z 247.0925. (c) Theoretical MS1 isotopic distribution for the proposed molecular formula $C_9H_{15}N_2O_6$ ($C_9H_{14}N_2O_6^+$ [M+H]$^+$), simulated at a resolving power of 35,000, shown for comparison with the experimental spectrum. (d) MS2 fragmentation spectrum of the precursor ion m/z 247.0925 at collision energy 70 eV. Fragment ions are annotated with their elemental compositions (mass error <3 ppm). The nitrogen-free ions at m/z 115.0389 ($C_5H_7O_3^+$), 97.0284 ($C_5H_5O_2^+$), 85.0284 ($C_4H_5O_2^+$) and 69.0336 ($C_4H_5O^+$) are consistent with a pentose moiety. Ions at m/z 157.0607 ($C_6H_9N_2O_3^+$) and 111.0552 ($C_5H_7N_2O^+$) retain both nitrogen atoms.

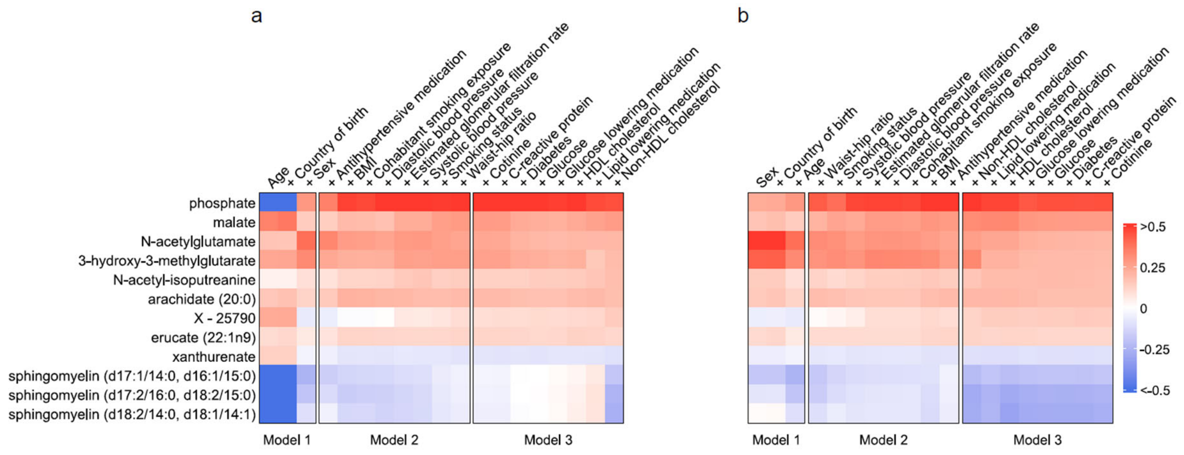


**Supplementary Figure 2.** Associations of coronary atherosclerosis-associated metabolites in models adding one covariate at the time where a) covariates are added within each model in alphabetical order, or b) covariates are added within each model in reverse alphabetical order. The colors of the heatmap indicate the strength and direction of the regression coefficients after addition of the covariate on top of each column. BMI: body mass index; HDL: high-density lipoprotein; SIS: segment involvement score.

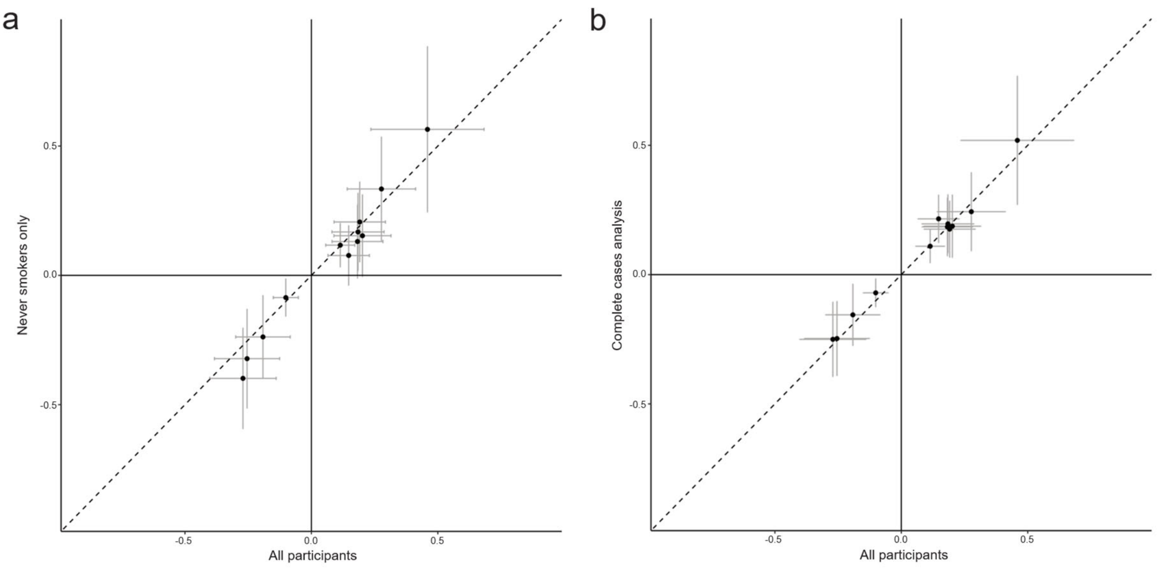


**Supplementary Figure 3.** Sensitivity analysis comparing results for coronary atherosclerosis-associated metabolites from Model 3 when **a)** excluding both current and former smokers (n=4,139), or **b)** using complete cases (n=6,489), with the main analysis including all participants (n=8,146). Regression coefficients with the 95% confidence intervals were plotted. The dashed diagonal line is where values of y = x. The horizontal line depicts y=0, and the vertical line x=0.

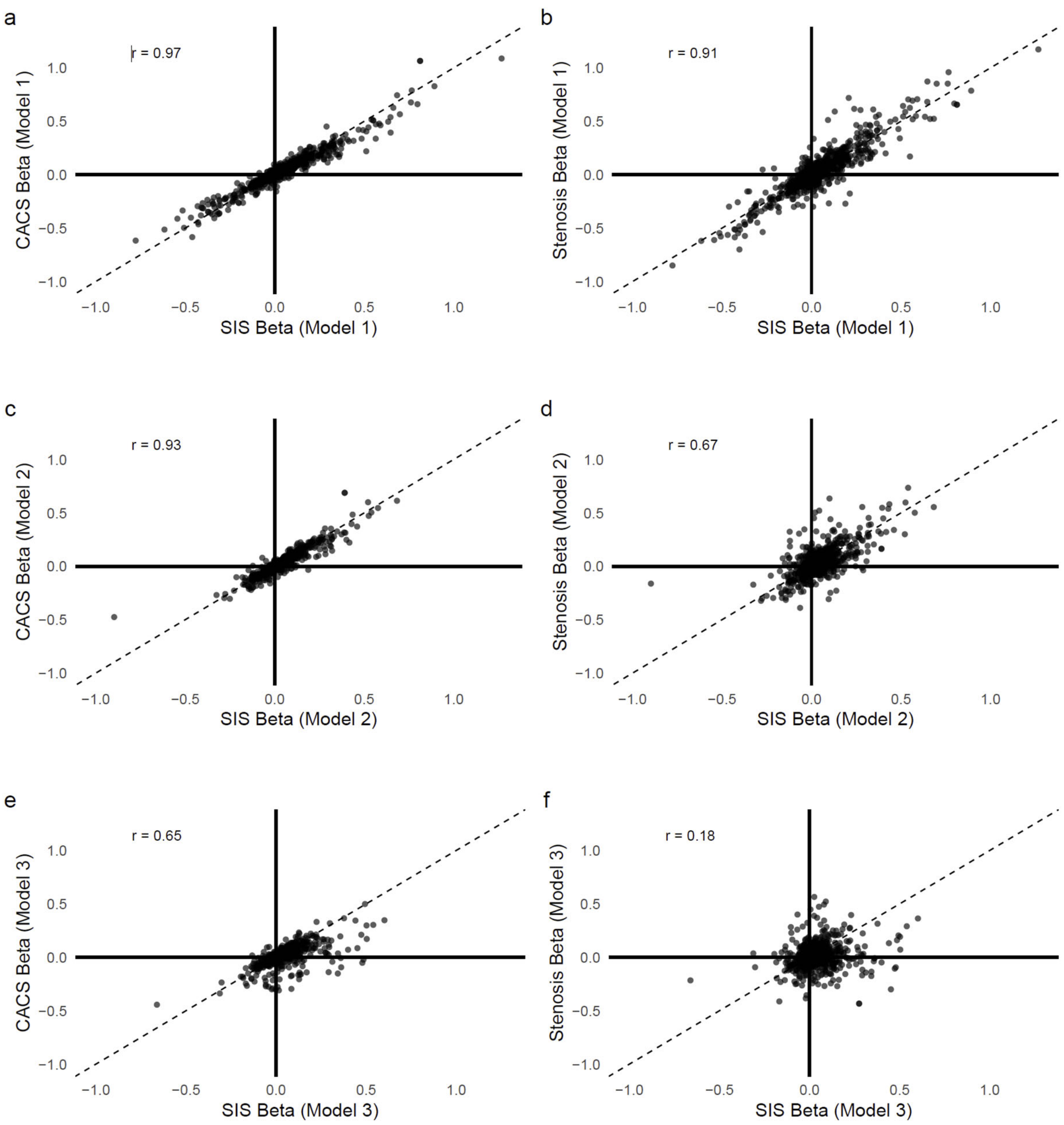


**Supplementary Figure 4.** Comparison of regression coefficients for metabolites associated with segment involvement score (SIS) with those associated with coronary artery calcium score (CACS) and ≥50% stenosis. Displayed are correlations for SIS and a) coronary artery calcium score (CACS) in Model 1, b) ≥50% stenosis in Model 1, c) CACS in Model 2, d) ≥50% stenosis in Model 2, e) CACS in Model 3, and f) ≥50% stenosis in Model 3. The dashed diagonal line is where values of y=x. Pearson correlation coefficients between the compared values are shown on the top left corner of each panel. Model 1 was adjusted for age, sex, and country of birth. Model 2 included additional adjustments for smoking status, anthropometric traits, estimated glomerular filtration rate (eGFR), blood pressure, and use of antihypertensive medication. Model 3 was further adjusted for lipid levels, glycemic markers, use of lipid-lowering and antidiabetic medications, C-reactive protein, and cotinine.

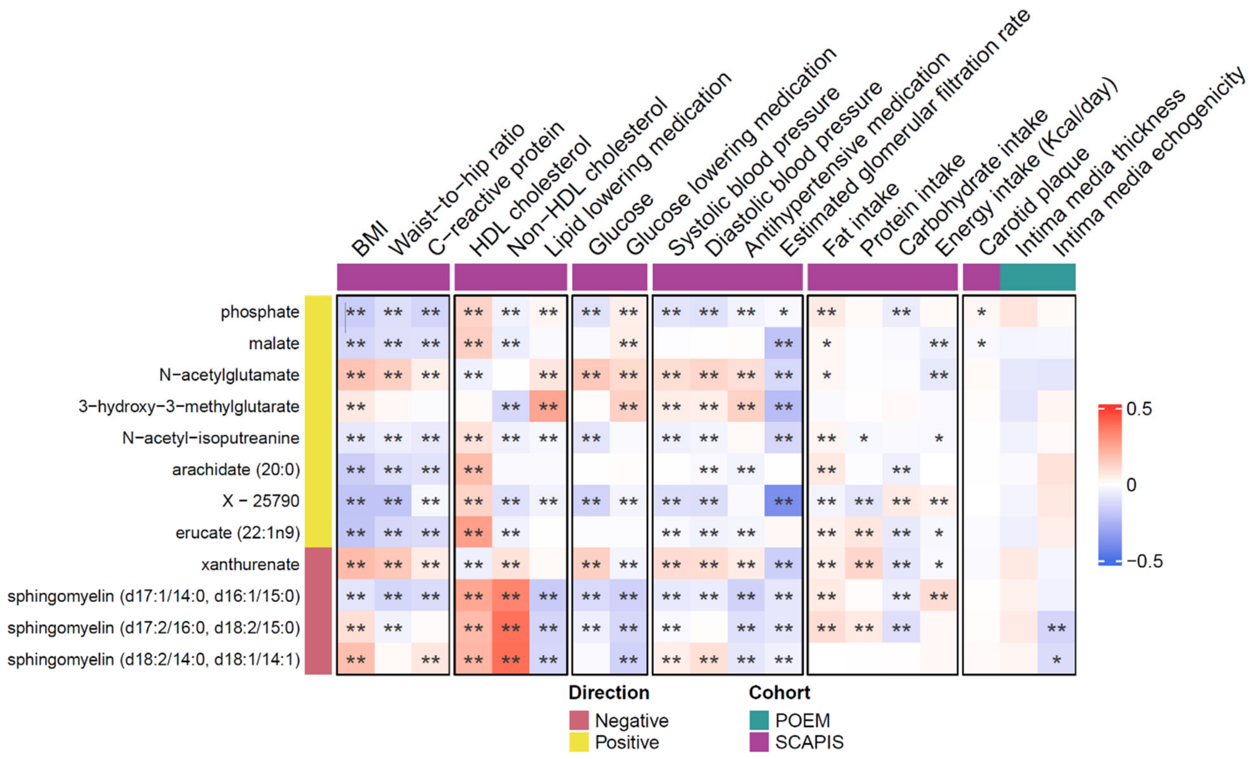


**Supplementary Figure 5.** Partial Spearman correlations of coronary atherosclerosis-associated metabolites with cardiovascular risk factors and medications adjusted for age, sex and country of birth in the SCAPIS participants without cardiovascular disease, and with carotid phenotypes in SCAPIS (n=8,146) and POEM (n=487). Direction of association between metabolites and SIS in Model 3. The color intensity reflects the magnitude of the unscaled correlation between the corresponding pairs of atherosclerosis-associated metabolites and cardiovascular risk factors and medications. Statistical significance: ** FDR $q < 0.05$; * nominal $p < 0.05$ but FDR-adjusted $q \geq 0.05$; no stars: nominal $p \geq 0.05$. BMI: body mass index; FDR: false discovery rate; HDL: high-density lipoprotein; POEM: Prospective Investigation of Obesity, Energy and Metabolism; SCAPIS: Swedish CArdioPulmonary bioImage Study; SIS: segment involvement score.
